# Ex-vivo 7T MRI for accurate margin assessment in oral squamous cell carcinoma surgery

**DOI:** 10.64898/2026.10.12.26365284

**Authors:** Jamila Guichelaar, Rémi van der Woude, Klijs de Koning, Jan Willem Dankbaar, Gerben Breimer, Annette van der Toorn, Rob Noorlag, Remco de Bree, Marielle Philippens

## Abstract

**Introduction:** Resection margins in oral squamous cell carcinoma (OSCC) are frequently inadequate. This study investigated the accuracy and diagnostic performance of ex-vivo 7T MRI against histology.

**Methods:** Fifty OSCC specimens underwent ex-vivo 7T MRI. Tumours were segmented on corresponding histology and twice on MRI: blinded to histology (radiologist segmentation; clinical practice) and using histology (histology-MR segmentation; MR-based ground truth). Resection margins were measured in five regions and compared with histology.

**Results:** Anterior and posterior margins were generally smaller on MRI than on histology (maximum mean difference -2.5 mm), whereas cranial, caudal and deep margins were generally larger on MRI (mean differences <1 mm). For close margins (<5 mm), sensitivity was 95% for histology-MR segmentations and 91% for radiologist segmentations, with 75% specificity for both.

**Conclusion:** Ex-vivo 7T MRI accurately measures resection margins and detects close margins with high sensitivity. A future clinical trial will determine whether intraoperative implementation reduces inadequate margins and adjuvant treatment.

## Introduction

Oral squamous cell carcinoma (OSCC) is one of the most common head and neck cancers worldwide.[1,2] OSCC is primarily treated by surgical resection. The goal of surgery is to remove the tumour with adequate resection margins, as complete tumour removal is essential for locoregional control and disease-free survival.[3,4] However, in clinical practice, resection margins are frequently inadequate. Studies report inadequate (i.e., close and positive) margins in 30% to 85% of cases, especially in submucosal and deep soft tissues.[5–7] Inadequate margins are associated with higher rates of local recurrence, distant metastasis, and reduced overall survival, and are therefore the main indication for adjuvant treatment (i.e., re-resection or radiotherapy).[4,8] Both adjuvant treatment options carry serious risks and side-effects. Adjuvant radiotherapy can drastically reduce the patient’s quality of life due to significant morbidity, and (oral) discomfort, including xerostomia, mucositis, fibrosis, and osteoradionecrosis.[9] Reresection often requires general anaesthesia and introduces new surgical risks, along with challenges in accurately relocating the inadequate margin. Therefore, obtaining adequate resection margins during the initial surgery is crucial to minimise the need for adjuvant treatment.

During surgery, surgeons typically assess the resection margins by visually inspecting and palpating the tumour and excised tissue. In addition, a frozen section analysis (FSA) of the resection specimen can be performed. However, FSA has low sensitivity in detecting close or positive margins due to sampling and interpretation errors.[10–12] Consequently, there is a clear need for alternative methods to improve intraoperative margin assessment. Over the past decade, several imaging modalities have been studied, and research continues to expand in this field.[13] Recent studies have investigated the application of intraoperative ultrasound (US).[14,15] Although the implementation of US has led to more adequate margins, several limitations remain. Firstly, the interpretation of US can be challenging in areas of the oral cavity with multiple tissue layers, the presence of salivary glands, and bony structures, such as the mandible and maxilla. Secondly, spatial orientation can be difficult, as the US images are in 2D, have a limited field of view, and lack spatial references.[14,16]

An imaging modality that could overcome these limitations is ex-vivo Magnetic Resonance Imaging (MRI). Ex-vivo MRI can be implemented for all oral cavity tumours, provides good soft tissue contrast, and allows intraoperative visualisation of resection margins in multiple directions.[17–21] In our centre, ex-vivo MRI is performed using a preclinical 7 Tesla (T) small-bore system with high gradient performance, enabling high spatial resolution imaging within a clinically reasonable acquisition time. In this study, we evaluate the accuracy of this 7T MRI system in detecting inadequate resection margins in OSCC resection specimens. We compare margin measurements on ex-vivo MRI to corresponding histological measurements as a reference standard. Furthermore, we investigate the clinical performance of ex-vivo MRI by comparing the radiologists’ assessments of margin status with the histological outcome.

## Methods

### Patients

Patients with OSCC scheduled for surgical tumour resection were prospectively included between July 2022 and April 2025. The study was approved by the medical research ethics committee (approval ID: 20-788), and all patients provided written informed consent prior to surgery.

### Specimen Preparations

Immediately after surgical excision, each specimen was marked with sutures to preserve anatomical orientation. Specimens were prepared for MRI and histological processing using a standardised workflow to ensure identical orientation across both modalities (Figure 1).

**Figure 1:**
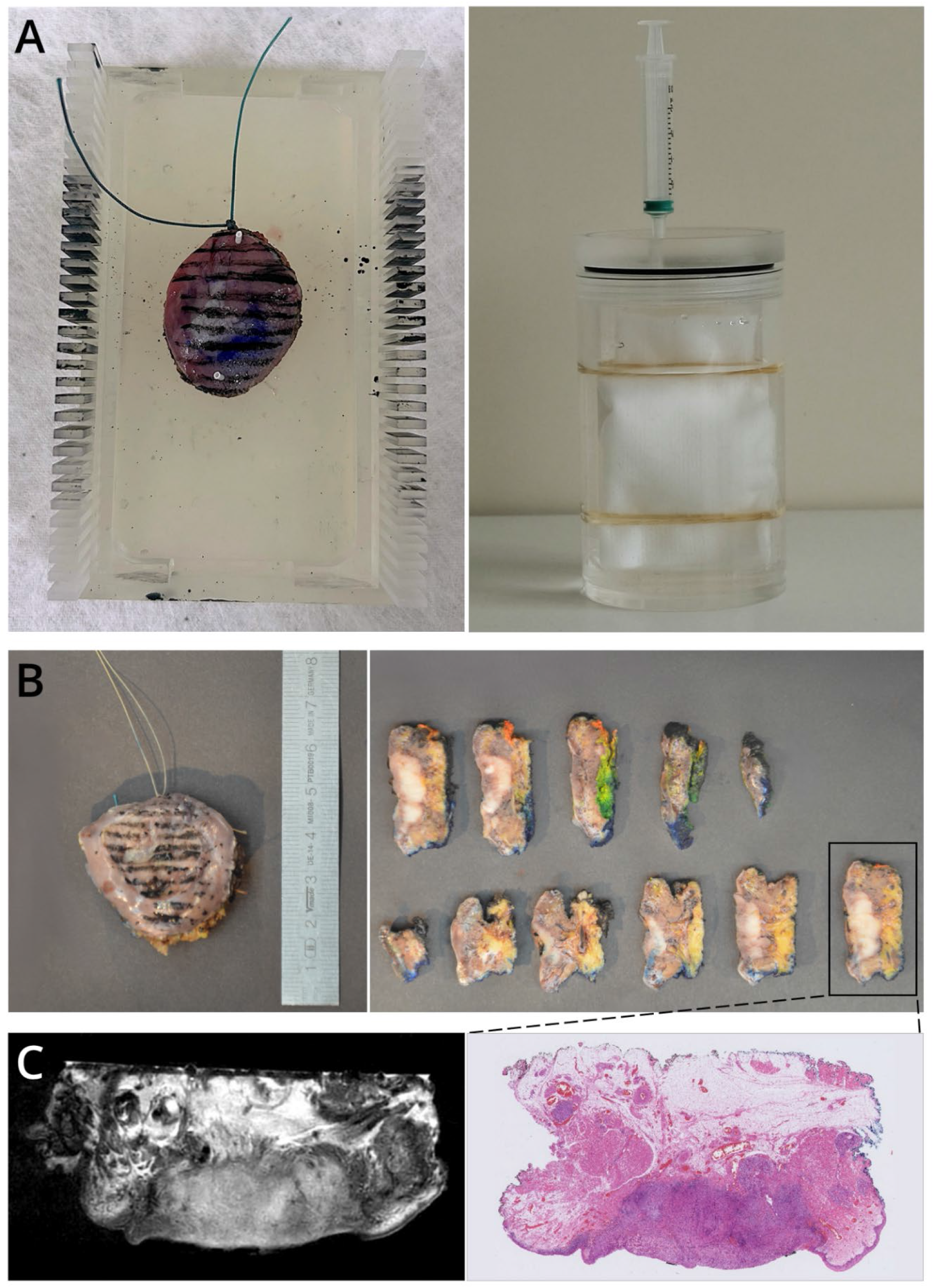
Standardised workflow for ex-vivo 7T MRI and histological processing of OSCC specimens. A) Left: freshly resected tumour specimen secured with cactus needles on the custom specimen tray inside the 3D-printed sectioning guide. Right: specimen secured to the specimen tray using a gauze and elastic bands and submerged in PFPE within the cylinder for ex-vivo MRI. B) Fixed specimen with black sectioning lines (left) and the resulting 3-mm thick sections (right), with one section highlighted with a black box to indicate the corresponding HE-stained slice in panel C. C) Example of an ex-vivo 7T MR slice (left) and the corresponding histological HE-stained slice (right).

Each specimen was pinned onto a silicone mat using cactus needles[22] and placed on a custom-made specimen tray. The tray was positioned in a 3D-printed sectioning guide, containing evenly spaced notches to standardise the orientation of the histological cuts. Using these notches, black ink lines were applied to the mucosal surface. After MRI, these lines guided the pathologist to section the specimen in the same orientation as the MR images.

The specimen was secured to the tray using a gauze and two elastic bands and placed in a polymethyl-methacrylate (PMMA) cylinder (outer diameter: 70 mm, inner diameter: 59 mm). This PMMA cylinder was then filled with PFPE (Galden, Solvay Solexis, Thorofare, NJ, USA) to minimise air-tissue susceptibility artefacts during MRI.

### MRI Acquisition

All specimens were scanned immediately after preparation using a small-bore 7T MRI scanner (BioSpec 7T, Bruker, Ettlingen, Germany), with a 0.66 T/m gradient system and operated using a Philips console (Philips Medical Systems, Best, The Netherlands).

The specimen-containing cylinder was positioned inside a quadrature transmit-receive volume coil (inner diameter: 72 mm; Bruker, Ettlingen, Germany) with the mucosal surface facing down to ensure that the specimen was fully submerged in PFPE.

The B_0_ field was homogenised using Compass software (Tesla Dynamic Coils, Zaltbommel, Netherlands) with second-order image-based shimming. A transverse and sagittal T2-weighted (T2w) Turbo Spin Echo (TSE) sequence was acquired with an in-plane resolution of 0.125 x 0.125 mm^2^, and slice thickness of 0.9 mm (Table 1). For small specimens, the echo train length was reduced from 15 to 5 at a fixed echo time (TE) to improve the signal-to-noise ratio (SNR) by reducing the voxel bandwidth, while maintaining clinically acceptable scan times.

**Table 1:**
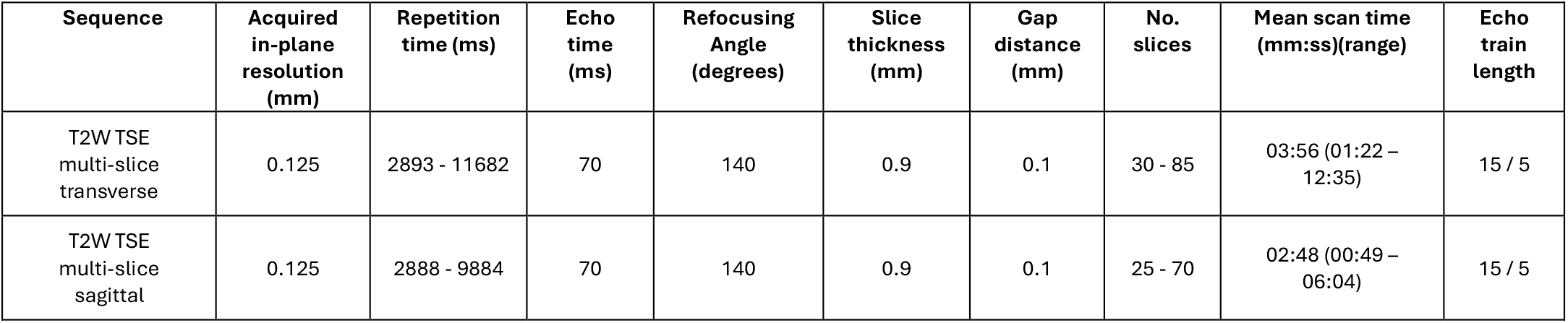
Sequence parameters of ex-vivo 7T MRI.

| Sequence | Acquired in-plane resolution (mm) | Repetition time (ms) | Echo time (ms) | Refocusing Angle (degrees) | Slice thickness (mm) | Gap distance (mm) | No. slices | Mean scan time (mm:ss)(range) | Echo train length |
| --- | --- | --- | --- | --- | --- | --- | --- | --- | --- |
| T2W TSE multi-slice transverse | 0.125 | 2893 - 11682 | 70 | 140 | 0.9 | 0.1 | 30 - 85 | 03:56 (01:22 – 12:35) | 15 / 5 |
| T2W TSE multi-slice sagittal | 0.125 | 2888 - 9884 | 70 | 140 | 0.9 | 0.1 | 25 - 70 | 02:48 (00:49 – 06:04) | 15 / 5 |

### Histology

Following MRI, specimens underwent routine histological processing. Specimens were fixed in 4% formaldehyde solution for a minimum of 24 hours. Subsequently, specimens were cut along the previously applied lines into ∼3-mm sections. The sections were transferred to standard or whole-mount cassettes for paraffin embedding. Of each section, a 4-µm slice was obtained and stained with haematoxylin and eosin (HE). All HE-stained slices were digitised at 40x magnification using Hamamatsu NanoZoomer S360 and S60 scanners (Hamamatsu Photonics K.K., Hamamatsu City, Japan)[23,24].

### Tumour Segmentation

Tumours were segmented on the digitised histological slices and MR images by two observers (RvdW and CJG). To prevent the histopathology results from influencing MR segmentations, the observers alternated modalities for each specimen. For each specimen, one observer performed the MR segmentation, while the other performed segmentation on the histological slices. This ensured that neither observer segmented both modalities for the same specimen.

MR segmentations were created using dedicated software (Volumetool, version V1.30.60)[25]. The tumour was segmented primarily on transverse T2w images and refined on sagittal T2w images when necessary. All MR segmentations were reviewed and corrected by an experienced head-and-neck radiologist (JWD; >10 years experience), who was blinded to the histopathology results. The resulting MR segmentations are hereafter referred to as “radiologist segmentations”. All histopathological segmentations were reviewed and corrected by an experienced head-and-neck pathologist (GB; >6 years experience). These segmentations are hereafter referred to as “pathologist segmentations”.

### Registration

Each histological slice was visually matched to the corresponding MR slice with the highest visual resemblance in Volumetool. Affine point-based registration of paired histological and MR slices was performed using at least three corresponding anatomical landmarks, such as tumour protrusions, mucosa edges, or specimen contours (Supplementary Material 1). Registrations were manually adjusted when necessary.

The registered histological slices were digitally stacked to create a 3D-volumetric representation of the histological sections[26]. The pathologist segmentations were propagated to the transverse MR scan using the transformation matrix derived from the point-based registration. After propagation, the pathologist segmentations were slightly manually adjusted to correct for deformation or tearing during histological processing and will now be referred to as the “histology-MR segmentation”.

### Measurements

The tumour depth of invasion (DOI) and resection margins were measured on histological slices and MR images. Re-resections attached to the specimen were not included in the measurements. The measurements followed the method described by Bekedam et al.[27], which subdivides the resection plane into deep, cranial, and caudal regions. A reference line was drawn between the two ends of the mucosal surface. From this line, at the middle of the tumour, two lines were drawn at 45° angles relative to the reference line. The region enclosed by the two 45° lines was defined as the deep margin, whereas the outside regions were defined as the cranial or caudal regions, depending on specimen orientation (Figure 2).

**Figure 2:**
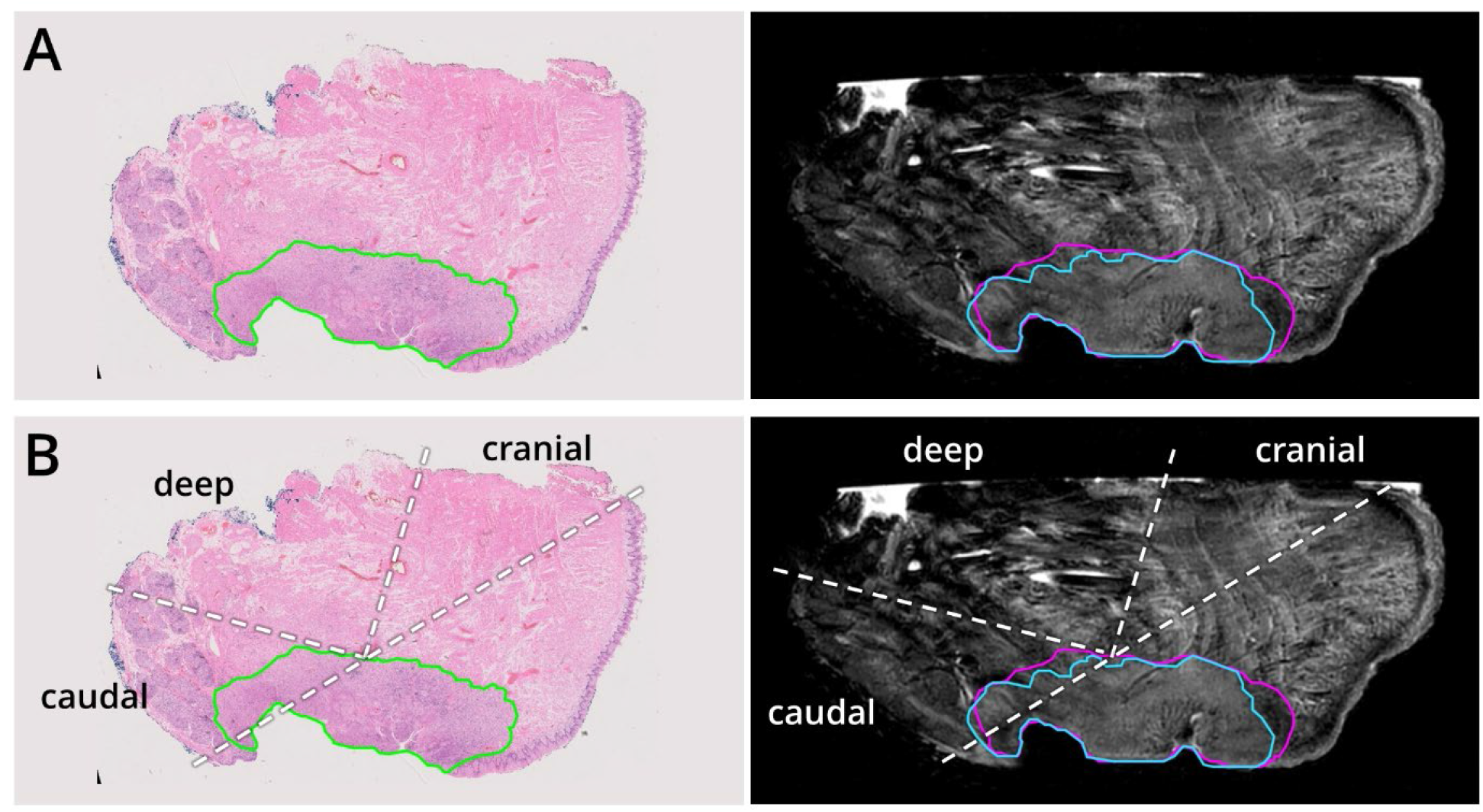
Workflow for measuring resection margins. A) Paired histological slice (left) and ex-vivo MR slice (right) shown with the three corresponding tumour segmentations: pathologist segmentation (green), radiologist segmentation (blue), and histology-MR segmentation (purple). B) Illustration of the measurement regions, where the dotted white lines divide the resection plane into the caudal, deep, and cranial regions.

Measurements for the anterior and posterior resection margins differed between histology and the MRI. On histology, these margins were estimated by counting the number of tumour-free slices and multiplying by the section thickness. On MRI, anterior and posterior margins were measured directly on sagittal T2w images.

For each specimen, the closest margin was measured in all five regions (i.e., anterior, posterior, cranial, caudal, and deep). Three sets of resection margin measurements were performed for each specimen: one set of histological margins measured on histology using the pathologist segmentation, and two sets of MR margins measured on the ex-vivo MR images using the histology-MR segmentation and radiologist segmentation. The radiologist margins reflect the assessment of ex-vivo MRI in clinical practice, whereas the histology-MR margins provide the closest MR-based ground truth.

### Analysis

All analyses were performed using IBM SPSS Statistics (Version 31.0; IBM Corp.).

#### Margin measurements

To evaluate the agreement between the reference histological and two MR margins, the difference between the margin measurements (MR margins minus histological margins) was calculated. The differences were calculated for each MR margin (based on histology-MR and radiologist segmentation) and for all five margin regions separately. The differences were plotted against the corresponding reference histological margin, and the mean and standard deviation (SD) of the differences were calculated. Upper and lower limits of agreement (LoA) were defined as the mean difference ±1.96 x SD. The same analysis was performed for the DOI measurements.

#### Diagnostic performance

To investigate the diagnostic performance of ex-vivo MRI for predicting margin status, a receiver operating characteristic (ROC) curve analysis was performed. In this analysis, the closest MR margin per specimen was compared with the final histological margin status. Two margin statuses were evaluated: (1) close margins, defined as <5 mm on histology, and (2) positive margins, defined as <1 mm on histology. If the closest margin on MRI was located at bone or periosteum (often <1 mm), this margin was excluded, since these bone margins may not be considered positive in clinical practice. In these cases, the next closest soft-tissue margin was considered the closest MR margin.

The ROC curve analysis was performed separately for the two MR margins and both histological margin statuses. Additionally, the area under the curve (AUC) with 95% confidence intervals (CI), sensitivity, and specificity were calculated.

## Results

A total of 68 patients were included. After excluding 18 resection specimens due to size constraints (n=4), metal or dental implants (n=4), MRI artefacts (n=2), histological artefacts (n=6), and logistical issues (n=2), the final cohort consisted of 50 patients. Patient characteristics are presented in Table 2.

**Table 2:** Patient Characteristics.

| Characteristics | n |
| --- | --- |
| *Total patients* | 50 |
| Males | 24 (48%) |
| Females | 26 (52%) |
| *Age (years)* |  |
| Mean (SD) | 63 (±13) |
| Range | 33 – 87 |
| *Tumour location* |  |
| Tongue | 27 (54%) |
| Buccal mucosa | 14 (28%) |
| Floor of mouth | 6 (12%) |
| Retromolar trigone | 2 (4%) |
| Gingiva | 1 (2%) |
| *pT stage* |  |
| pT1 | 13 (26%) |
| pT2 | 24 (48%) |
| pT3 | 9 (18%) |
| pT4a | 4 (8%) |
| *Tumour depth of invasion (mm)* |  |
| Mean (SD) | 7.4 (±4.9) |
| Range | 1.0 – 21.3 |
| *Worst pattern of invasion (WPOI)* |  |
| 1 | 0 |
| 2 | 5 (10%) |
| 3 | 10 (20%) |
| 4 | 20 (40%) |
| 5 | 15 (30%) |
| *Tumour occurrence* |  |
| Primary | 37 (74%) |
| Second primary | 13 (26%) |
| *Histological margin status\** |  |
| Free (≥5 mm) | 8 (16%) |
| Close (1-5 mm) | 30 (60%) |
| Positive (<1 mm) | 12 (24%) |

\* Bone margins and intraoperative reresections not included.

### Margin measurements

The differences between MR and histological margins varied across the five margin regions. For the anterior and posterior margins, the mean differences were negative, indicating smaller margins on MRI than on histology. The cranial, caudal and deep margins all showed a positive mean difference, indicating larger MR margins than the corresponding histological margin. The scatter plots of the caudal and deep margin are shown in Figure 3. The plots of the remaining three regions are provided in Supplementary Material 2. In addition to the overall trends, a few specimen-level discrepancies between histological and MR margin measurements were observed (Figure 4).

**Figure 3:**
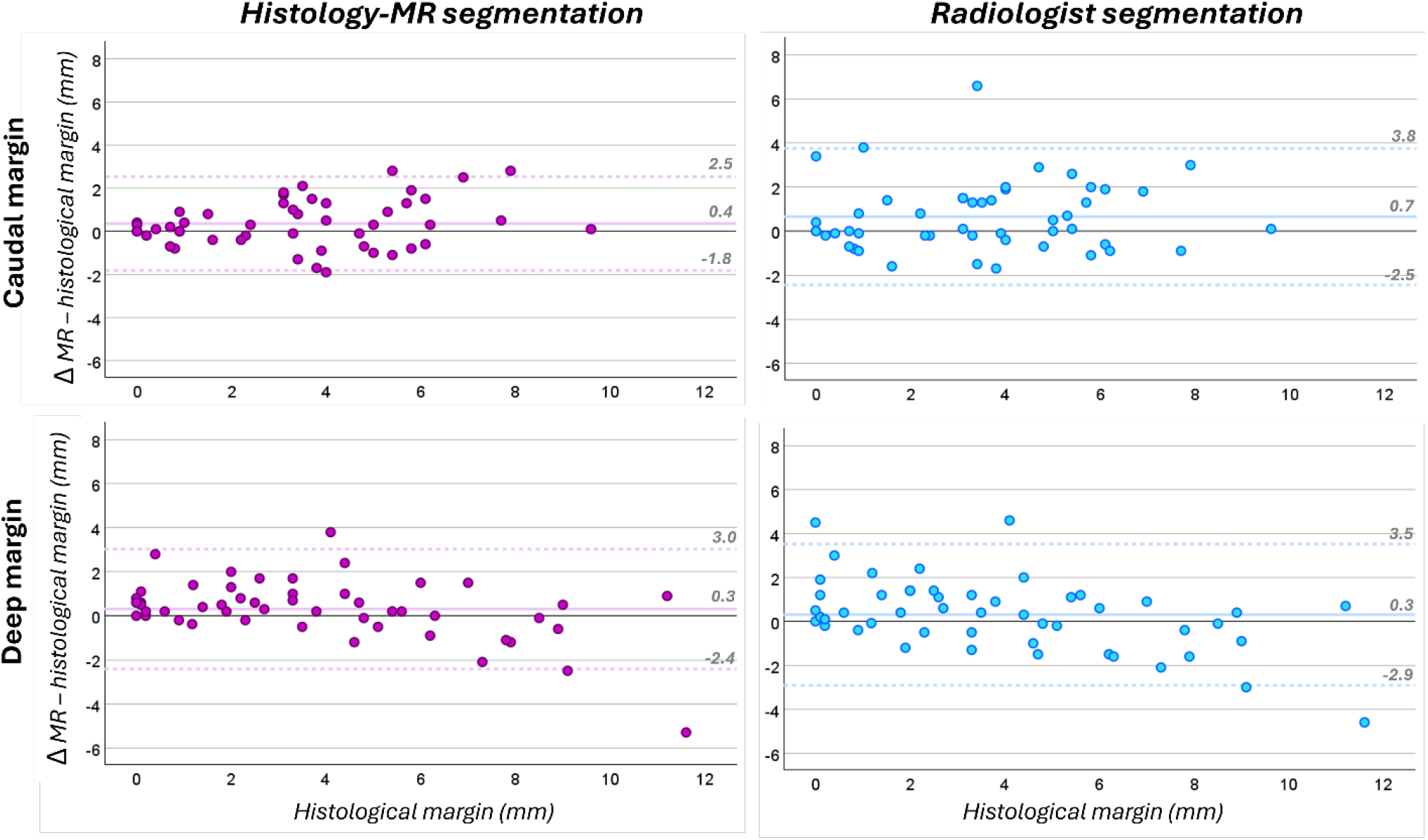
Difference between MR and histological margins in the deep and caudal region. The differences are plotted against the histological reference margin and shown separately for each MR margin (histology-MR segmentation in purple, radiologist segmentation in blue). Each point corresponds to one specimen. The mean difference is indicated with a solid horizontal line. The dashed horizontal lines indicate the upper and lower LoA. The differences for the anterior, posterior and cranial regions are shown in Supplementary Material 2.

**Figure 4:**
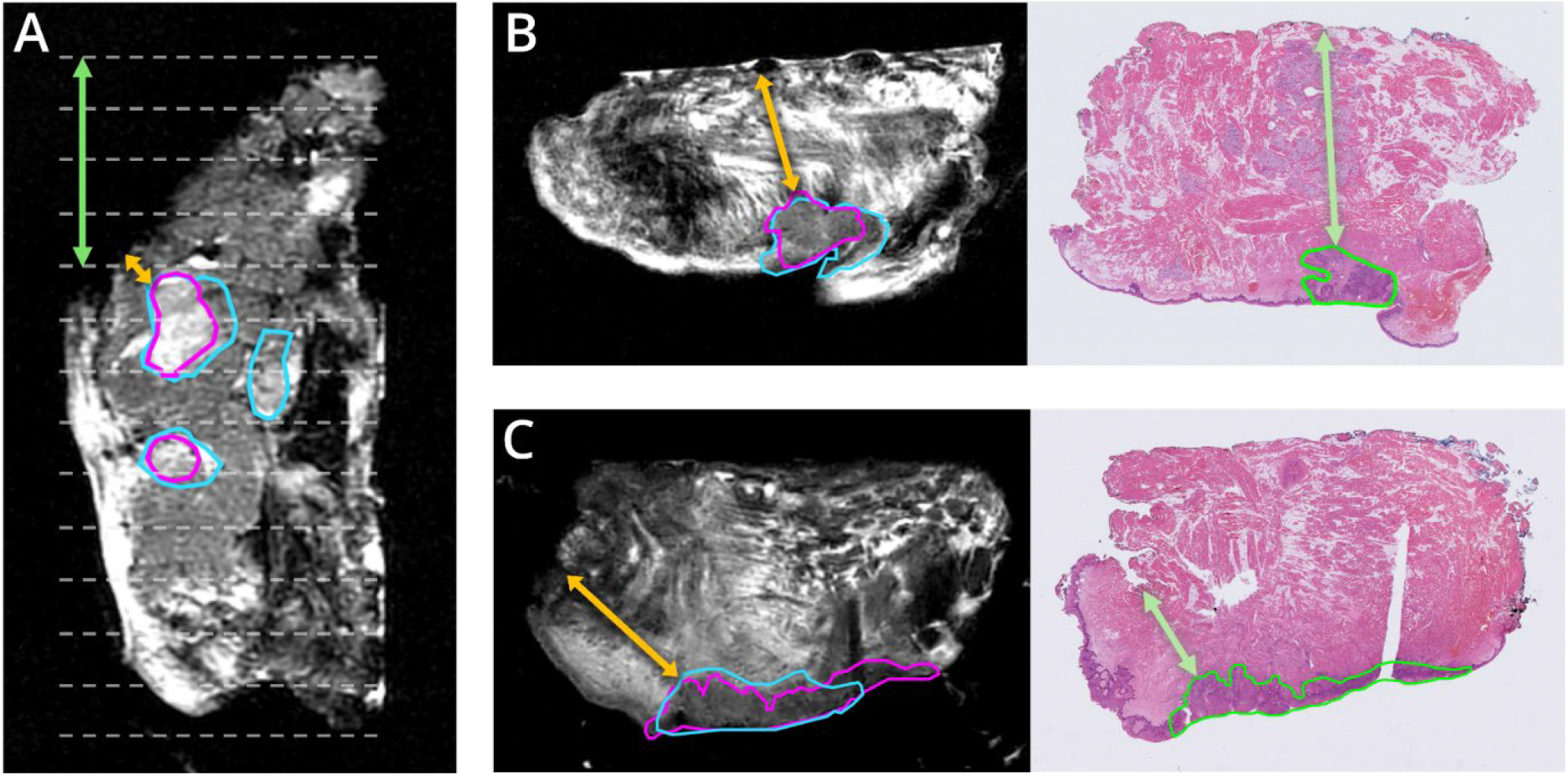
Discrepancies between margin measurements on histology and MRI. The pathologist segmentation is indicated in green, histology-MR segmentation in purple, and radiologist segmentation in blue. The green arrow represents the histological margin, and the orange arrow represents both MR margins. A) Sagittal MR image demonstrating an anterior margin difference due to the different measuring methods. The white dashed lines indicate the histological sections. The histological margin measures 4 tumour-free slices, while a significantly smaller margin is measured on MRI. B) Deep margin difference caused by deformation of the specimen due to compression against the specimen tray during the MRI acquisition. C) Margin discrepancy due to an unclear specimen contour. The histological slice shows irregularities along the resection plane that are difficult to distinguish on MRI.

For the histology-MR segmentation, the anterior and posterior margins showed mean differences of -2.5 mm (LoA: -11.5 to 6.5 mm) and -0.2 mm (LoA: -6.9 to 6.4 mm), respectively. The cranial margin had a mean difference of 0.5 mm (LoA: -1.8 to 2.7 mm), the caudal margin 0.4 mm (LoA: -1.8 to 2.5 mm), and the deep margin 0.3 mm (LoA: -2.4 to 3.0 mm).

For the radiologist segmentation, the anterior and posterior margins showed mean differences of -1.8 mm (LoA: -11.6 to 8.0 mm) and -0.4 mm (LoA: -8.3 to 7.5 mm), respectively. The cranial margin had a mean difference of 0.7 mm (LoA: -2.8 to 4.3 mm), the caudal margin 0.7 mm (LoA: -2.5 to 3.8 mm), and the deep margin 0.3 mm (LoA: -2.9 to 3.5 mm).

### DOI measurements

On average, DOI measurements were larger on MRI than on histology. For the histology-MR segmentation, the mean difference to the histological DOI was 0.8 mm (LoA: -1.3 to 2.8 mm). For the radiologist segmentation, the mean difference was 1.0 mm (LoA: -2.0 to 4.1 mm). The scatter plots of the DOI differences are provided in Supplementary Material 3.

### Diagnostic performance

According to histology, 42 of the 50 specimens had close resection margins, of which 12 had positive margins <1 mm.

In general, ex-vivo MRI showed good diagnostic performance in detecting histological margins <5 mm. For the histology-MR segmentation, the AUC was 0.96 (95% CI: 0.91–1.01). When using a 5-mm cut-off on MRI, the sensitivity and specificity were 95% and 75%, respectively. For the radiologist segmentation, the AUC was 0.94 (95% CI: 0.88–1.01), with a sensitivity of 91% and specificity of 75% (Figure 5).

**Figure 5:**
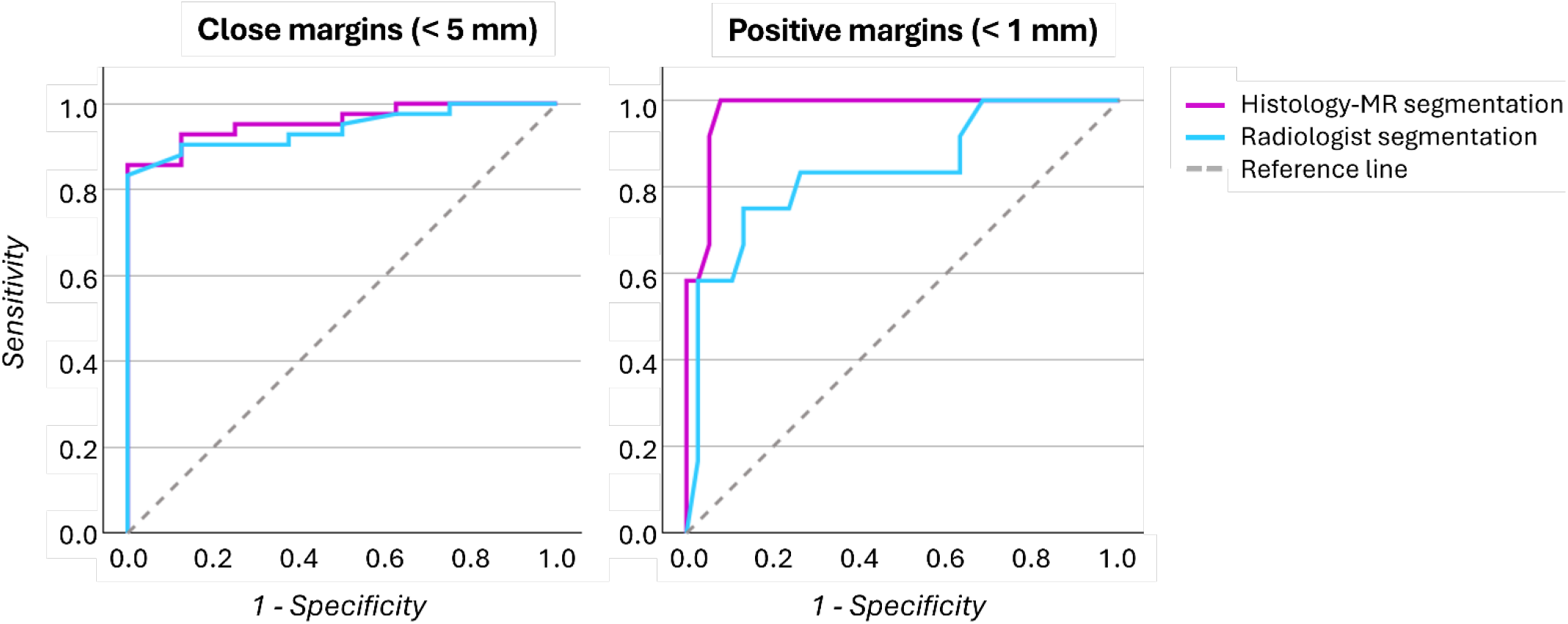
ROC curve analysis. Diagnostic performance of ex-vivo MRI in detecting close margins (left) and positive margins (right). The analysis was performed for each MR margin separately (histology-MR segmentation in purple, radiologist segmentation in blue). The diagonal reference line represents chance-level performance.

For detecting positive resection margins, the AUC of the histology-MR segmentation was 0.98 (95% CI: 0.94–1.01). When using a 1-mm cut-off on MRI, the sensitivity and specificity were 83% and 95%, respectively. The radiologist segmentation showed an AUC of 0.84 (95% CI: 0.70–0.98), with a sensitivity of 58% and specificity of 97% (Figure 5).

## Discussion

In this study, we evaluated the accuracy of ex-vivo 7T MRI in detecting inadequate resection margins in OSCC surgery. The ROC analysis showed high sensitivity and high AUC values for detecting both close and positive margins when margins were measured using the histology-MR segmentation. Using the radiologist segmentation, which more closely reflects clinical practice, close margins were also predicted with high sensitivity. However, the sensitivity for detecting margins <1 mm was moderate. This lower performance is partly explained by the small number of specimens with positive margins in our cohort (12/50). This makes sensitivity estimates less stable, since a few misclassifications can substantially affect the performance metrics. Furthermore, the 1-mm threshold defines a very narrow classification range, making cases susceptible to misclassification even when differences between MRI and histology are small. This was also reflected in our data: five of the 12 specimens were misclassified as >1 mm based on the radiologist segmentation, while two of these had margins slightly above the 1-mm threshold (1.6 and 1.3 mm on MRI compared to 0.9 and 0.8 mm on histology). The other three specimens were classified as having close margins. It is important to note that these false negatives do not necessarily imply that the positive margins would have been missed in clinical practice, as immediate reresections would be indicated for margins below 5 mm.

When comparing margin measurements across the five regions, we found that MR margins were generally smaller than those on histology in the anterior and posterior regions, and larger in the cranial, caudal and deep regions. The differences in the anterior and posterior regions are most likely explained by the different measuring methods. On MRI, we measured the margins directly on the sagittal MR images, while the histological anterior and posterior margins are estimated by multiplying the number of tumour-free slices by the section thickness. One of the outliers in our data (Figure 4A) clearly demonstrates this issue: based on the tumour-free slices, the histological margin appears adequate, while MRI reveals a much smaller submucosal margin.

The cranial, caudal and deep margins provide a more reliable comparison between MRI and histology, as these margins were measured in the same transverse slices. Overall, both MR margins corresponded closely to histological margins, with small differences consistent with expected tissue shrinkage during histological processing.[28–31] Greater variability was observed for the radiologist segmentation, likely reflecting the combined effects of tissue shrinkage and inaccuracies in tumour segmentation. Nevertheless, margin assessment based on the radiologist segmentation still demonstrated high sensitivity for detecting close margins. Furthermore, the margin differences still remained below 0.8 mm, with deep margins being most accurate compared to histology (mean difference of 0.3 mm). This finding is encouraging, as deep margins are most often reported as inadequate.[5–7] We expect that MRI measures the deep margins most accurately because the closest deep margin is often located at the largest tumour bulk, which is clearly visible on MRI. Another explanation may be that slight compression of the specimen during MRI acquisition reduces the MR deep margin. This makes the margin more similar to the histological margin, which is inherently smaller due to tissue shrinkage. Combining the results of the ROC analysis and margin measurements, we believe that MRI has strong potential for detecting inadequate resection margins and improving margin assessment in OSCC surgery.

This is the first study to investigate ex-vivo MRI in a large and diverse cohort of OSCC specimens. A comparable study by Steens et al.[21] explored ex-vivo MRI in ten tongue SCC specimens using a similar 7T small-bore MRI system. They reported that small, superficial tumours were difficult to detect on MRI, consistent with our observations. For visible tumours, Steens et al. found that margins could be predicted within a 2 mm difference compared with histology. The improved performance observed in our study is likely attributed to higher scan quality. The smaller volume coil increased SNR by positioning the specimen closer to the coil. Additionally, a longer echo time (70 ms) and smaller voxel size enhanced T2-weighting and spatial resolution, respectively. This resulted in better tumour–tissue differentiation than by Steens et al.

As a follow-up to Steens et al., Heidkamp et al. explored the use of a 3T clinical MRI scanner to overcome the practical difficulties associated with the 7T MRI system[20]. While tumours could be correctly detected, accurate assessment of resection margins remained limited. This poor performance is most likely due to the changeover to a lower field strength and the resulting SNR loss, limiting visualisation of fine details. The high number of pT1 tumours in their cohort (5/10) likely further contributed to this poor performance, as these tumours are inherently more difficult to detect. Consequently, the findings of these studies suggest that the potential of ex-vivo MRI for intraoperative margin assessment is strongest when using a high-resolution 7T system.

One of the main strengths of this study is our large and diverse cohort of OSCC specimens, which allowed us to evaluate ex-vivo MRI for different tumour locations, sizes and characteristics. Additionally, our optimised MRI-protocol produced high-resolution T2-weighted images with good tissue contrast, clear tumour definition, and clinically reasonable acquisition times. Furthermore, our systematic workflow for registering the histological slices to MRI enabled accurate comparison with histology and to validate our results.

Besides these strengths, there are also limitations to be considered. First, the segmentations and margin measurements were performed in a postoperative, non-clinical setting. As a result, observers had time to segment tumours carefully and measure margins with high precision. When implementing ex-vivo MRI during surgery, margin assessment will likely be less precise due to time constraints. Therefore, the results in this study might overestimate the performance of ex-vivo MRI intraoperatively. Second, although we optimised the registration of histology to MRI by manual adjustments, some cases still showed clear mismatches due to tissue deformation or shrinkage, which may have affected some margin comparisons (Figure 4B).

Other limitations relate to tumour characteristics and MRI-specific factors. For instance, although image quality was adequate for most tumours, small or superficial (pT1) tumours were generally difficult to detect. As these tumours are often adequately resected, the added clinical value of ex-vivo MRI is limited in this group. Additionally, in some cases, the transition between the mucosa and resection plane was not clearly visible on MRI, making it difficult to assess submucosal margins. Furthermore, the exact specimen contour was not always well defined, especially along bone or where the resection plane was irregular or torn. In some cases, this led to significantly larger margins on MRI than on histology (Figure 4C). In clinical practice, these issues may be mitigated by assessing pre-acquired photographs of the specimen during margin assessment.

A final limitation of ex-vivo 7T MRI is the challenge of integrating it into the surgical workflow, as not all centres have access to a small-bore 7T MRI system. In our centre, the scanner is located in a separate department from the operating room, meaning that the entire workflow (including transfer to the scanner) can be time-consuming. However, we have optimised specimen preparation, scanning, and margin assessment, and can complete the total workflow within 45 minutes. This timeframe is comparable to FSA, which can be replaced by ex-vivo 7T MRI. In most OSCC surgeries, tumour resection is combined with treatment of the neck, either via a sentinel node procedure or an (elective) neck dissection. The ex-vivo 7T MRI workflow can be completed during treatment of the neck, enabling intraoperative margin assessment without extending overall surgical time.

Future research should focus on validating ex-vivo MRI in clinical practice. We are currently working towards a prospective clinical trial to implement intraoperative margin assessment with ex-vivo 7T MRI. The main objective will be to reduce the need for adjuvant treatment by lowering the amount of inadequate resection margins. Subsequently, we will assess whether ex-vivo 7T MRI will lead to better oral function and quality of life. We will exclude cT1 tumours due to limited detectability and expected clinical benefit.

To conclude, we demonstrated that ex-vivo 7T MRI can accurately predict inadequate resection margins with high sensitivity in OSCC specimens. Margins could be measured with high precision when compared to the histological reference standard. A future clinical trial will validate these findings in clinical practice.

## Data Availability

The data that support the findings of this study are not publicly available due to privacy and ethical restrictions concerning patient data.

## Acknowledgements

This work was supported by KWF, the Dutch Cancer Society, grant number: 14384. KWF funded this work but did not influence the research.

