## Supplementary Material for "Ex-vivo 7T MRI for accurate margin assessment in oral squamous cell carcinoma surgery"

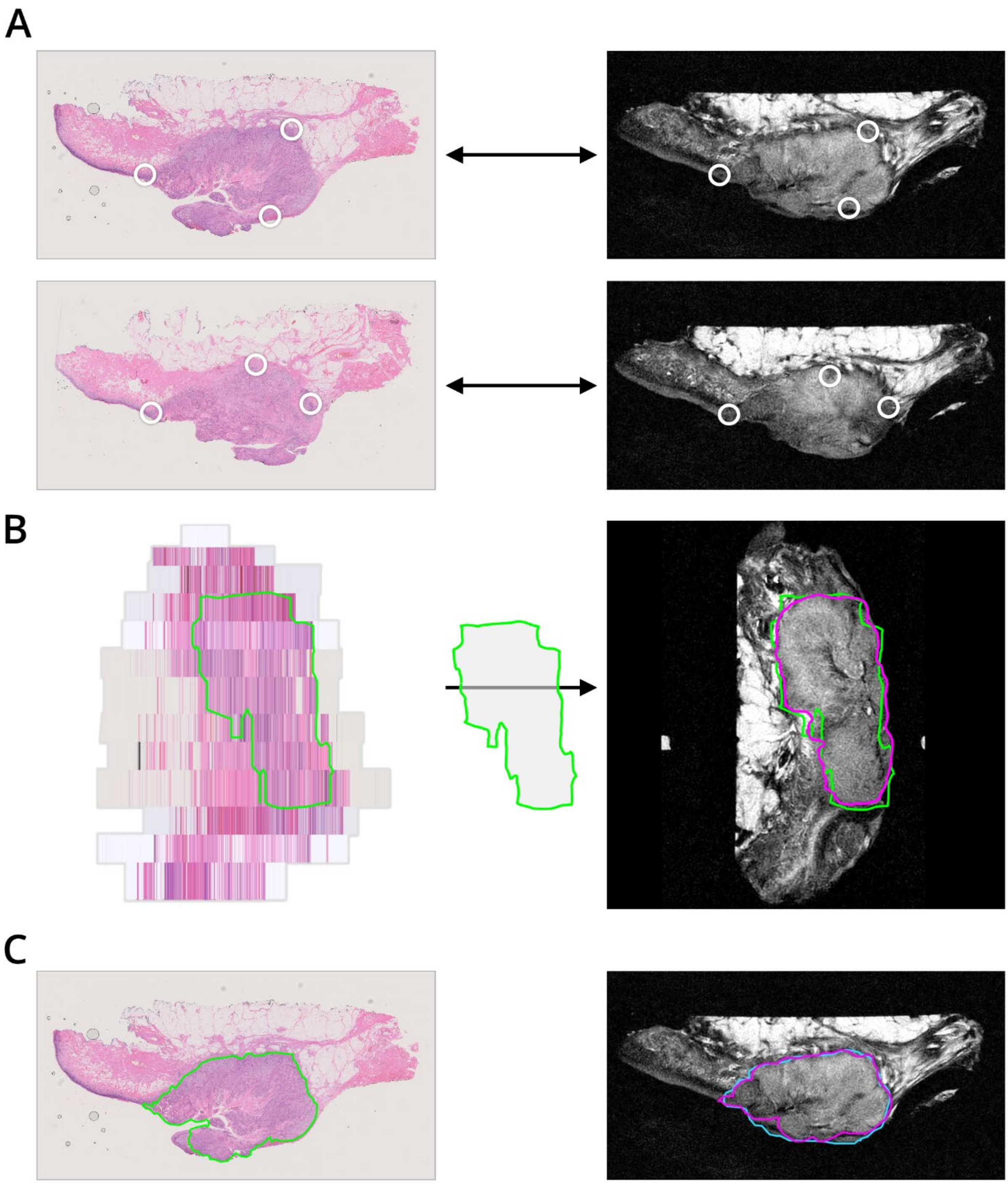


*Supplementary Material 1:* **Registration of histology data to ex-vivo 7T MRI.** A) Two histological slices (left) and their visually matched transverse MR slices (right), each annotated with at least three matching anatomical landmarks (white circles) used for affine point-based registration. B) All registered histological slices were digitally stacked to create a 3D volumetric reconstruction of the histological slices (left). The pathologist tumour segmentation (green) was propagated to ex-vivo MRI, and then slightly manually adjusted to match the ex-vivo MRI anatomy, resulting in the histology-MR segmentation (purple). A sagittal ex-vivo MR image with histology and histology-MR segmentation is shown on the right. C) Example of a paired histological slice with the pathologist segmentation (green, left) and the corresponding transverse ex-vivo MR slice with the radiologist segmentation (blue, right) and histology-MR segmentation (purple, right)

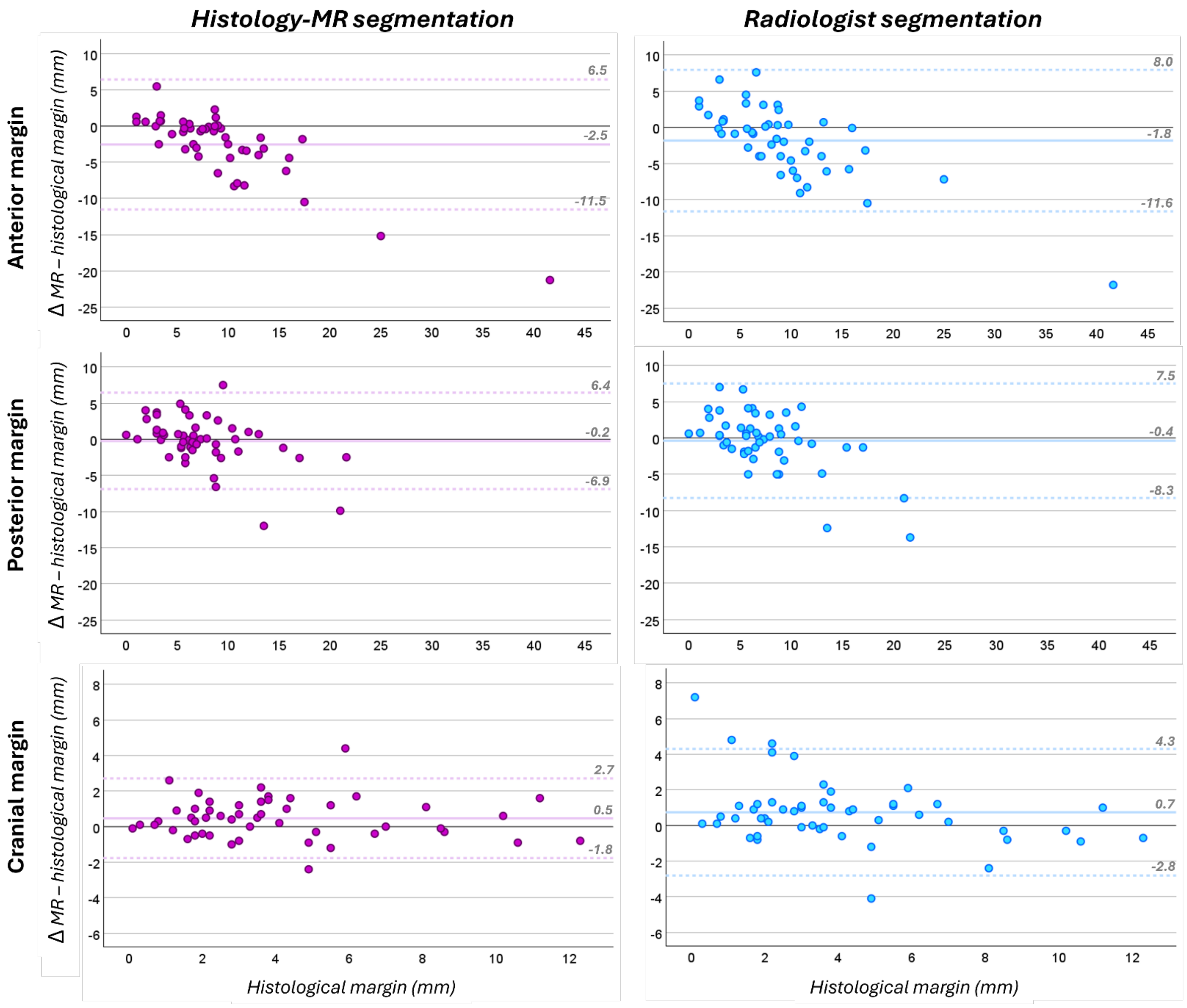


*Supplementary Material 2:* **Differences between MR and histological margins in the anterior, posterior, and cranial regions.** The differences are plotted against the histological reference margin and shown separately for each MR margin (histology-MR segmentation in purple; radiologist segmentation in blue). Each point corresponds to one specimen. The mean difference is indicated with a solid horizontal line, and the dashed horizontal lines indicate the upper and lower LoA.

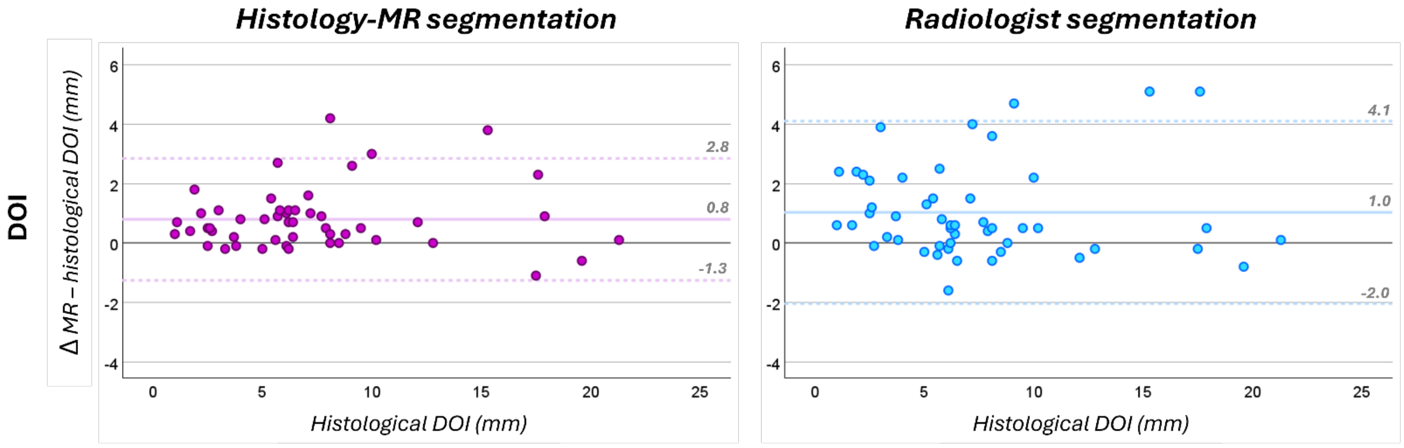


*Supplementary Material 3:* **Differences between MR and histological DOI.** The differences are plotted against the histological reference DOI and shown separately for each MR segmentation (histology-MR segmentation in purple; radiologist segmentation in blue). Each point corresponds to one specimen. The mean DOI is indicated with a solid horizontal line, and the dashed horizontal lines indicate the upper and lower LoA.
